# Systematic Testing for SARS-CoV-2 Infection Among Essential Workers in Montréal, Canada: A Prospective Observational and Cost Assessment Study

**DOI:** 10.1101/2021.05.12.21256956

**Authors:** Jonathon R. Campbell, Cynthia Dion, Aashna Uppal, Cedric P. Yansouni, Dick Menzies

**Affiliations:** Research Institute of the McGill University Health Centre, Montréal, Canada; Faculty of Medicine, McGill University, Montréal, Canada; McGill International TB Centre, Montréal, Canada; J.D. MacLean Centre for Tropical Diseases

**Keywords:** communicable diseases, prevalence, infection control, resource allocation, health services, health policy

## Abstract

**BACKGROUND:** Essential workers are at increased risk of severe acute respiratory syndrome coronavirus 2 (SARS-CoV-2). We did a prospective study to estimate the yield, acceptability, and costs of workplace-based systematic SARS-CoV-2 testing of asymptomatic essential workers.

**METHODS:** We recruited non-healthcare essential businesses, in Montréal, Canada. Mobile teams, composed of two non-healthcare professionals each, visited businesses. Consenting, asymptomatic employees provided saline gargle specimens under supervision. Mobile team members self-sampled weekly. Specimens were analyzed using reverse-transcription polymerase chain reaction (RT-PCR). If an outbreak was detected (≥2 positives), we retested all initially negative participants. We did logistic regression for factors associated with a positive test. We estimated costs ($CAD) of this strategy.

**RESULTS:** From 27 January to 12 March 2021, 69 essential businesses were visited. Of an estimated 2348 employees onsite, 2128 (90.6%) participated. Across 2626 tests, 53 (2.0%) were positive. Self-reported non-Caucasian ethnicity (aOR 3.7, 95% CI: 1.4-9.9) and a negative SARS-CoV-2 test before the study (0.4, 0.2-0.8) were positively and negatively associated with a positive test, respectively. Five businesses—3 manufacturing/supplier and 2 meat processing— were experiencing an outbreak. At these businesses, 40 (4.4%) of 917 participants were positive on the initial test. We repeated testing at three of these businesses over 2-3 weeks: 8/350 (2.3%) were positive on the second test, and zero were positive on the third and fourth test (148 tests); no employer reported new positives to 26 March 2021. In all other businesses, 1211 participants were tested once—5 (0.4%) were positive at three childcare enterprises, one grocery store, and one manufacturing/supplier. Per person, RT-PCR costs were $34.00 and all other costs $8.67. No mobile team member tested positive.

**INTERPRETATION:** Onsite sampling of essential workers with saline gargle is safe, acceptable, and inexpensive. Repeat testing appeared to eliminate outbreaks. Systematic testing should be considered part of SARS-CoV-2 preventive efforts.

## Background

Severe acute respiratory syndrome coronavirus 2 (SARS-CoV-2), responsible for coronavirus disease 2019 (COVID-19), is a public health emergency. Identification and isolation of infected persons is a key component of strategies to prevent and eliminate SARS-CoV-2.(1,2)

Essential workers are at increased risk of acquiring SARS-CoV-2.(3,4) While guidance is to stay home if symptomatic,(5) infected persons may be contagious despite having mild or non-specific symptoms(6) or no symptoms at all.(7,8) Frequent testing could identify asymptomatic infectious persons in workplaces and prevent further transmission. However, this not is not common practice. Instead, people with symptoms are encouraged to visit testing centres; this ignores transmission from asymptomatic persons plus means employees may need to take time off work, limiting uptake.(9) Additionally, the primary specimen collection method for SARS-CoV-2 is nasopharyngeal swabs.(10) These must be performed by a healthcare professional, require extensive personal protective equipment, are uncomfortable, and expensive.(9,11,12) Onsite sampling with a saliva-based method that does not require a healthcare professional would relieve financial and human resource demand and improve feasibility.(13)

We conducted a prospective study in Montréal, Canada to assess the yield, acceptability, and costs of workplace-based, systematic collection of saline gargle samples from asymptomatic essential workers for SARS-CoV-2 testing.

## Methods

### Setting

This study took place from January 27, 2021 through March 12, 2021, in Montréal, Canada, primarily within the borough of Montréal-North. Until January 26, 2021, the cumulative SARS-CoV-2 detection rate in Montréal-North was 8,088 per 100,000 population—nearly double the cumulative rate in Montréal overall (4,445 per 100,000 population).(14) The study immediately followed the peak of the second wave in Québec. During the study, the provincial weekly SARS-CoV-2 test positivity rate fell from 4.0% to 2.7%.(15)

Prior to study start, on December 25, 2020, non-essential businesses in Montréal were closed and a province wide curfew from 8pm to 5am was implemented on January 9, 2021. During the study period, on February 8, 2021, non-essential businesses were re-opened, including personal care services and shopping centres, and on February 26, 2021, movie theatres, swimming pools, and skating rinks were opened with some restriction.(16)

### Procedures

Before the study, we engaged with the mayor’s office, business representatives, and health authorities in the borough of Montréal-North, as well as Montréal Public and Occupational Health to refine study priorities and procedures. We assembled two mobile sample collection teams consisting of two members each. Mobile team members were not healthcare professionals; they received five days of training on research, study procedures, and infection control and prevention measures. Mobile team members were tested weekly during the study for SARS-CoV-2 infection.

Businesses deemed essential according to the Québec government were contacted to participate.(17) Businesses interested in participating provided an estimate of the number of employees that would be present on the day of testing for scheduling purposes.

On the day of testing, the mobile team(s) set up sampling stations at the business, which included a table with chairs placed on opposite sides and a plexiglass divider in the middle. Stations were sanitized before and after each participant. Mobile team members wore gloves (changed between participants) and at least one three-layer surgical mask but did not wear face shields/eye protection nor gowns. All participants had to wear masks (cloth or surgical) and sanitize their hands.

Eligible employees were at work that day, were at least 18 years old, and had not had a positive SARS-CoV-2 test within the past 4 weeks. To verify eligibility, all workers meeting the study team were asked “Do you have any symptoms that are unusual or out of the ordinary from how you feel any other day” (i.e., not associated with pre-existing conditions or their occupation) and COVID-19 testing in the past 4 weeks. Eligible workers were introduced to the study, provided signed informed consent, then completed an electronic questionnaire regarding demographic, clinical, and data on previous SARS-CoV-2 testing and illnesses, as well as current symptoms (Table e1). Questionnaire responses were reviewed post-visit. Under the supervision of a mobile team member, each participant provided a saline gargle sample to be tested for SARS-CoV-2. Participants placed 5ml of saline solution in their mouth, swished the solution for 5 seconds, gargled the solution for 5 seconds, and repeated the procedure; all liquid was placed in a tube and sealed (Appendix for procedures). Participants sanitized their hands and returned to work. Sample tubes were sanitized and packaged for transport to the lab.

Samples were analyzed following reverse-transcription polymerase chain reaction (RT-PCR) protocols by the laboratory.(18) Results were transmitted to the study team 4-24 hours later, who then notified participants immediately. Participants testing negative were notified by email or text message. Participants testing positive were notified by phone, asked to isolate (and not go to work), and provided information on social support available to them; subsequent management was done by Montréal Public Health.

If two or more positive participants were identified in a workplace, this met the definition of an outbreak and we notified Montréal Public and Occupational Health. We contacted the business to offer to return within 4-5 days to retest all initially negative employees and those who were not initially tested. Subsequently, we offered a third round of testing 4-5 days after our second visit and a fourth round of testing 14-21 days after our third visit. Montréal Occupational Health referred an additional three businesses who were at the onset of outbreaks to the study team. We followed identical procedures with these businesses.

### Cost Assessment

We estimated the unit costs of training, scheduling, sample collection, sample transport, RT-PCR, and contacting participants, using a health system perspective in 2021 Canadian dollars ($). We only considered costs that would be incurred outside of a research study using a microcosting approach (Table e2 for cost details). For capital costs (e.g., training, plexiglass) we assumed they would need to be repeated or replaced annually. To estimate costs of personnel time, we used time estimation questionnaires. We used the reimbursement cost in Québec for a sample undergoing laboratory RT-PCR.(19) We report costs per participant sampled, assuming a two-person team visited two 50-person businesses each day. We varied this assumption in sensitivity analysis. Additional details are in the Appendix.

### Data Analysis

To estimate a SARS-CoV-2 infection prevalence of 5% with 2% precision (95% CI 3% to 7%), we initially calculated a sample size of 2,589 participants would be required (see Appendix for details). We estimated the yield of testing overall (i.e., prevalence), and among different subgroups. We did logistic regression using generalized linear mixed-models to estimate adjusted odds ratios (aOR) and 95% confidence intervals (95% CI) for potential factors associated with a positive result.(20) We treated each business and sector as a random effect. We considered participant age (continuous), sex (male vs. female), self-reported ethnicity (Caucasian vs. non-Caucasian), health factor (reported any of hypertension, diabetes, respiratory condition, heart disease, or other health condition vs. not), smoking history (current or previous smoker vs. never smoker), feeling on day of testing as reported on electronic questionnaire (“fine” vs. “not my best today”), previous testing history (never previously tested vs. previously tested and always negative vs. previously tested and positive), and business size (≤50 participants vs. >50 participants). To deal with quasi-complete separation associated with some fixed effects, we used weakly informative priors.(21) For businesses experiencing an outbreak and where repeat testing was performed, we described the investigations and plotted the evolution of the outbreaks over time. Analysis was done in R (version 4.0.3) using base packages or package blme (version 1.0-5).

## Results

From January 27, 2021, and March 12, 2021, 69 businesses (19% of 366 contacted) were visited. Of 2348 eligible employees estimated onsite, 2138 (91%) visited the study team, and 2128 (90.6%) consented to participate (Figure 1). When asked by research staff, no participant mentioned “out of the ordinary” symptoms to the study team (i.e., all participants would classify themselves as asymptomatic). However, 30 (1%) participants reported not feeling their best on the questionnaire.

**Figure 1.**
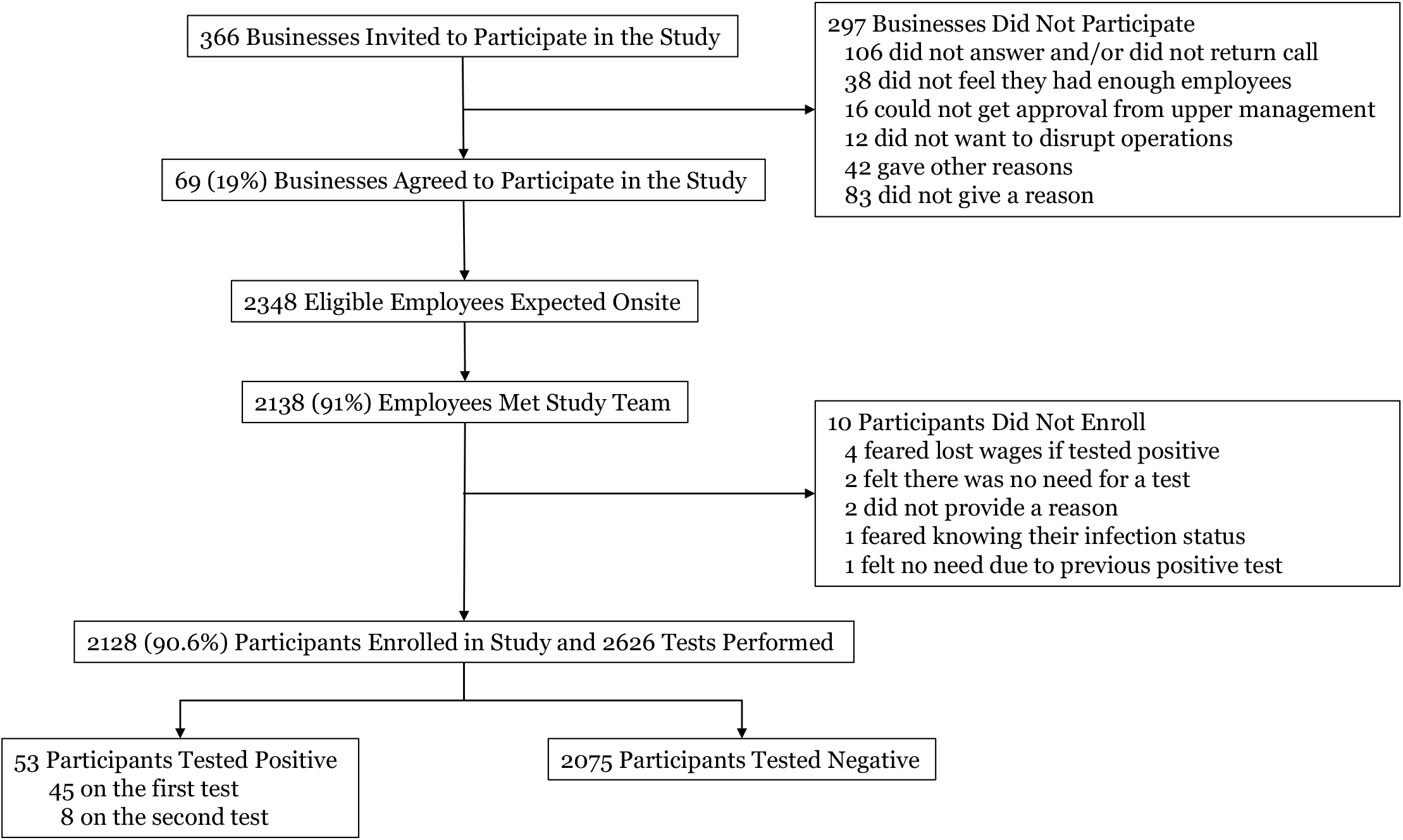
Flow diagram of participants included in the study and reasons for exclusion.

The median (IQR) number of participants per business was 13 (8 to 22). Most businesses (61; 88%) had ≤50 participants. Most participants (1225; 58%) worked in manufacturing or supplier sectors (Table 1). Among participants, 808 (38%) were female and the median (IQR) age was 48 (37 to 57) years. Overall, 973 (46%) participants were tested for SARS-CoV-2 at some point before the study—882 (41%) had never tested positive and 91 (4%) had tested positive more than 4 weeks prior to study enrollment (Table 2).

**Table 1.** Characteristics of included businesses

| Characteristic | Number of Businesses | Number of Participants |
| --- | --- | --- |
| **Total Businesses** | **69 (100%)** | **2128 (100%)** |
| Manufacturing/Suppliers | 19 (28%) | 1225 (58%) |
| Auto Sales and Repair | 12 (17%) | 242 (11%) |
| Childcare | 11 (16%) | 113 (5%) |
| Public Services | 8 (12%) | 144 (7%) |
| Grocery | 6 (9%) | 100 (5%) |
| Retail | 6 (9%) | 84 (4%) |
| Construction | 4 (6%) | 28 (1%) |
| Meat Processing | 2 (3%) | 183 (9%) |
| Legal | 1 (1%) | 9 (<1%) |
| **Median (IQR) Participants Per Business** | **13 (8 to 22)** | **--** |
| Size: 1-5 participants | 11 (16%) | 44 (2%) |
| Size: 6-10 participants | 14 (20%) | 113 (5%) |
| Size: 11-20 participants | 26 (38%) | 388 (18%) |
| Size: 21-50 participants | 10 (14%) | 350 (16%) |
| Size: 51 participants or more | 8 (12%) | 1233 (58%) |

Abbreviations: IQR, interquartile range

**Table 2.** Characteristics of included participants

| Characteristic | All Participants<br>N (%) | Participants Testing Positive<br>N (%) | Participants Testing Negative<br>N (%) |
| --- | --- | --- | --- |
| **Total Participants** | 2128 | 53 (2.5%) | 2075 (97.5%) |
| **Median (IQR) Age** | 48 (37 to 57) | 49 (40 to 58) | 48 (37 to 57) |
| **Sex** |  |  |  |
| Female | 808 (38%) | 9 (17%) | 799 (39%) |
| Male | 1320 (62%) | 44 (83%) | 1276 (61%) |
| **Self-Reported Ethnicity** |  |  |  |
| Caucasian | 926 (44%) | 4 (8%) | 922 (44%) |
| Non-Caucasian | 1202 (56%) | 49 (92%) | 1153 (56%) |
| Asian | 444 (21%) | 17 (32%) | 427 (21%) |
| Black | 295 (14%) | 21 (40%) | 274 (13%) |
| Hispanic | 204 (10%) | 6 (11%) | 198 (10%) |
| Indigenous | 4 (0%) | 0 (0%) | 4 (0%) |
| Mixed | 17 (1%) | 0 (0%) | 17 (1%) |
| Other/Not Disclosed | 238 (11%) | 5 (9%) | 233 (11%) |
| **Employment Status** |  |  |  |
| Full-Time Employee | 2018 (95%) | 51 (96%) | 1967 (95%) |
| Part-Time or Occasional Employee | 110 (5%) | 2 (4%) | 108 (5%) |
| **Reported Health Factor** |  |  |  |
| None | 1681 (79%) | 45 (85%) | 1636 (79%) |
| Any* | 447 (21%) | 8 (15%) | 439 (21%) |
| Hypertension | 214 (10%) | 4 (8%) | 210 (10%) |
| Diabetes | 151 (7%) | 4 (8%) | 147 (7%) |
| Chronic Respiratory Condition | 64 (3%) | 1 (2%) | 63 (3%) |
| Heart Disease | 31 (1%) | 0 (0%) | 31 (1%) |
| Other | 99 (5%) | 1 (2%) | 98 (5%) |
| **Reported Smoking** |  |  |  |
| Never Smoker | 1668 (78%) | 46 (87%) | 1622 (78%) |
| Current Smoker | 331 (16%) | 5 (9%) | 326 (16%) |
| Former Smoker | 129 (6%) | 2 (4%) | 127 (6%) |
| **Previous SARS-CoV-2 Test History\*\*** |  |  |  |
| Never Tested | 1155 (54%) | 42 (79%) | 1113 (54%) |
| All Previous Tests Negative | 882 (41%) | 9 (17%) | 873 (42%) |
| Any Previous Test Positive | 91 (4%) | 2 (4%) | 89 (4%) |
| **Feeling on the Day of Testing\*\*\*** |  |  |  |
| “Fine, Same as Any Other Day” | 2098 (99%) | 51 (96%) | 2047 (99%) |
| “Not My Best Today” | 30 (1%) | 2 (4%) | 28 (1%) |
| **Business Sector** |  |  |  |
| Manufacturing/Supplier | 1225 (58%) | 43 (81%) | 1182 (57%) |
| Meat Processing | 183 (9%) | 6 (11%) | 177 (9%) |
| Childcare | 113 (5%) | 3 (6%) | 110 (5%) |
| Other | 607 (29%) | 1 (2%) | 606 (29%) |
| **Business Size** |  |  |  |
| Size: 1-5 participants | 44 (2%) | 1 (2%) | 43 (2%) |
| Size: 6-10 participants | 113 (5%) | 2 (4%) | 111 (5%) |
| Size: 11-20 participants | 388 (18%) | 1 (2%) | 387 (19%) |
| Size: 21-50 participants | 350 (16%) | 0 (0%) | 350 (17%) |
| Size: 51 participants or more | 1233 (58%) | 49 (92%) | 1184 (57%) |

Abbreviations: IQR, interquartile range; SARS-CoV-2, severe acute respiratory syndrome coronavirus-2
\*Participants can have multiple health factors; \*\*refers to testing done prior to enrollment in the study; \*\*\*refers to reporting on electronic questionnaire (not to study team), which was reviewed post-visit
Note: percentages represent **column percentages**

We performed 2626 tests among the participants and 53 (2.0%) were positive. On their first test, 45 (2.1%) of 2128 participants in eight businesses tested positive for SARS-CoV-2. We performed subsequent testing at three businesses experiencing an outbreak. An additional 8 (2.3%) of 350 participants tested positive on their second test after testing negative on their first test. Of 121 and 27 participants who were tested a third and fourth time, respectively, none tested positive (Table e3). No research staff member tested positive during the study.

Among the 53 participants testing positive at any point during the study, 43 (81%) were in three different manufacturing and supplier businesses, 6 (11%) were in one meat processing facility, 3 (6%) were in three different childcare enterprises, and 1 (2%) was in a grocery store. Nearly all (49/53, 92%) of positive participants worked in businesses with >50 participants. A positive test result was more likely among those self-identifying as non-Caucasian (aOR: 3.7, 95% CI: 1.4 to 9.9) and less likely in those who had had at least one negative SARS-CoV-2 tests prior to study enrolment (aOR: 0.4, 95% CI: 0.2 to 0.8) (Table 3; Table e4 for unadjusted estimates).

**Table 3.**
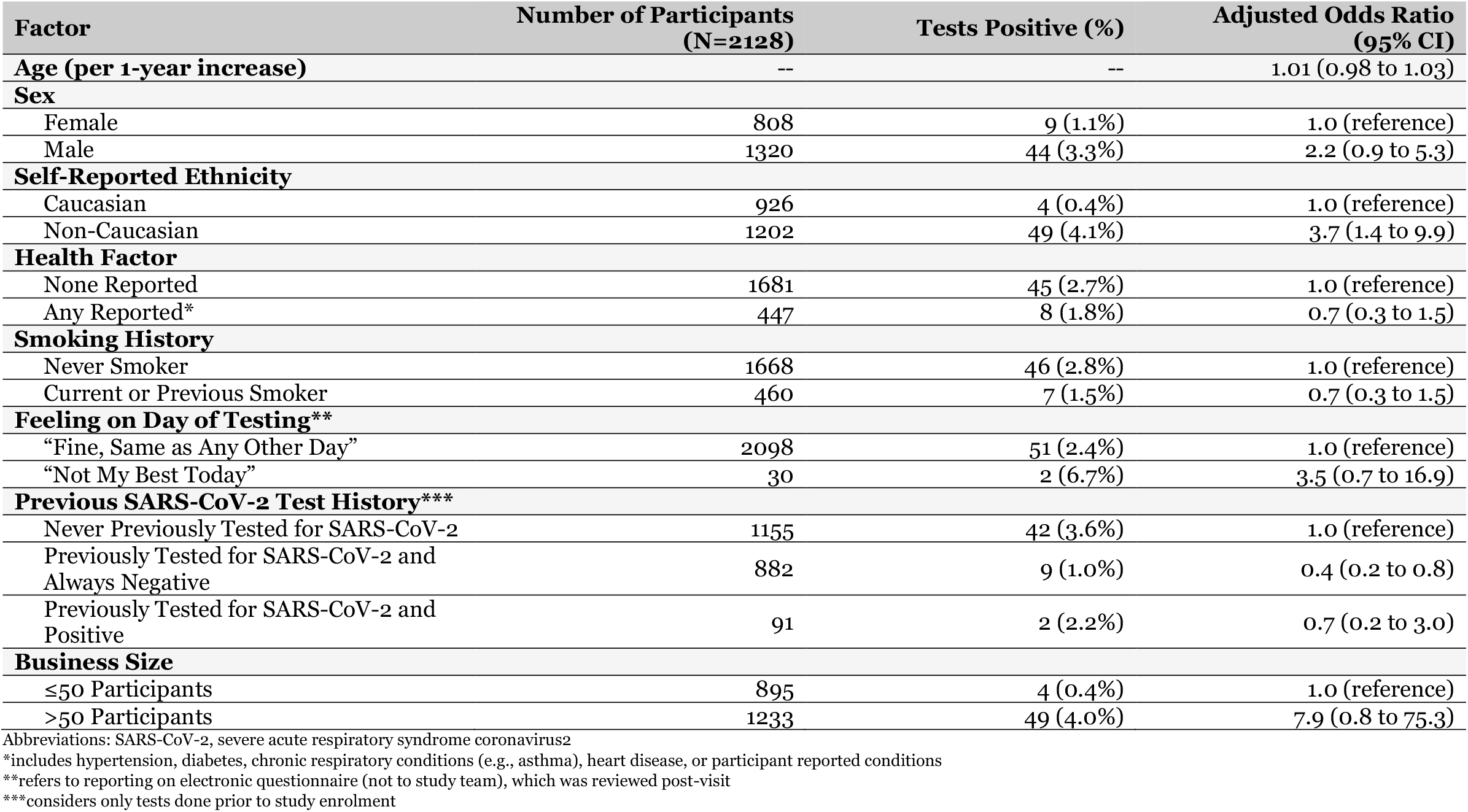
Logistic regression results for factors potentially associated with ever testing positive in the study. The model accounts for clustering by business and sector.

Among the 69 businesses visited, five businesses were experiencing an outbreak. In these businesses, 917 participants were tested at least once, of whom 40 (4.4%) were positive on initial testing. In businesses not experiencing an outbreak 5 (0.4%) of 1211 participants tested were positive. These infections occurred in three childcare enterprises, one manufacturing and supplier business, and one grocery store.

The total cost per person sampled was $42.67 (Table 4), of which $34.00 (80%) was for RT-PCR and $8.67 (20%) for all other costs. Personnel costs were lower if larger businesses were visited (Table e5).

**Table 4.** Cost ($ CAD) of training, scheduling, onsite saline gargle collection, transport, and contacting of participants per person sampled, assuming two 50-person businesses are visited per day by two mobile team members.

| Cost Estimates | Cost Per Person Sampled (\$ CAD) | Additional Details |
| --- | --- | --- |
| **TOTAL COST** | **\$42.67** | Sum of laboratory and non-laboratory costs |
| **TOTAL LABORATORY COST** | **\$34.00** | Reimbursement cost for RT-PCR to detect SARS-CoV-2 in Québec |
| **TOTAL NON-LABORATORY COST** | **\$8.67** | Sum of below components of training, scheduling, sample collection, sample transport, and communicating results |
| **Training of Mobile Team** | **\$0.04** | |
| Coordinator & Mobile Team Personnel | \$0.03 | Based on two days of training [estimated during study for non-research training days] |
| Materials & Refreshments | \$0.01 | Includes meals and materials for sample collection, sanitizing, and personal protective equipment* |
| **Scheduling Businesses** | **\$0.80** | |
| Administrative Personnel | \$0.80 | Based on two hours per business [estimated during study] |
| **Sample Collection** | **\$5.62** | |
| Mobile Team Personnel | \$2.99 | Based on 45 minutes to set up and tear down at each business and 6 minutes per person sampled [estimated during study] |
| Sanitizing Materials | \$0.33 | Includes rags, paper towels, disinfectant, and hand sanitizer* |
| Personal Protective Equipment | \$0.38 | Includes 4 masks per day and 2 gloves per participant* |
| Sample Collection Materials | \$1.74 | Includes a sample tube, a 5ml saline ampoule, 2 biohazard bags* |
| Capital Purchases | \$0.18 | Includes items such as chairs, tables, trash bins, plexiglass dividers, stationery, electronic data collection devices, storage bins, dollies, and coolers* |
| **Sample Transport** | **\$0.54** | |
| Mobile Team Personnel | \$0.19 | Assuming 30 minutes to drop off samples and 10 minutes driving between businesses [estimated during study] |
| Vehicle & Fuel | \$0.35 | Assuming 60km driven per day [estimated during study] |
| **Communicating Results** | **\$1.67** | |
| Administrative Personnel | \$1.67 | Based on 5 minutes per result on average [estimated during study] |

Abbreviations: SARS-CoV-2, severe acute respiratory syndrome coronavirus-2.
Key assumptions: one two-person mobile team can collect samples from 500 persons per week (100 per day); training would need to be repeated annually and capital purchases replaced annually—total costs for each of these were pro-rated over a year (i.e., divided by 26,000 persons sampled); coordinator salary of \$30/h, mobile team salary of \$23/h, administrative salary of \$20/h.
\*see Table e2, for more details.

### Details of SARS-CoV-2 Outbreaks

Two outbreaks were discovered serendipitously by the study team (Business A and B). In both instances, the employer then informed us that multiple symptomatic infections had been detected 3-4 weeks prior to our visit. Three were referred by public health (Business C, D, and E).

As shown in Figure 2A, over the course of four visits to Business A, test positivity fell steadily to reach 0% on the final visit. There were 4/39 (10.3%) and 1/68 (1.5%) conversions (negative-to-positive) on the second and third visit, respectively. Similar trends were observed in Business B (Figure 2B), where test positivity fell to 0% by the third round of testing; with 3/65 (4.6%) conversions detected on the second visit. Acceptance of repeat testing was 100% at Business A and 86% at Business B. Up to March 26, 2021, four weeks after our final visit to Business A and two weeks after our final visit to Business B, no new infections were reported.

**Figure 2.**
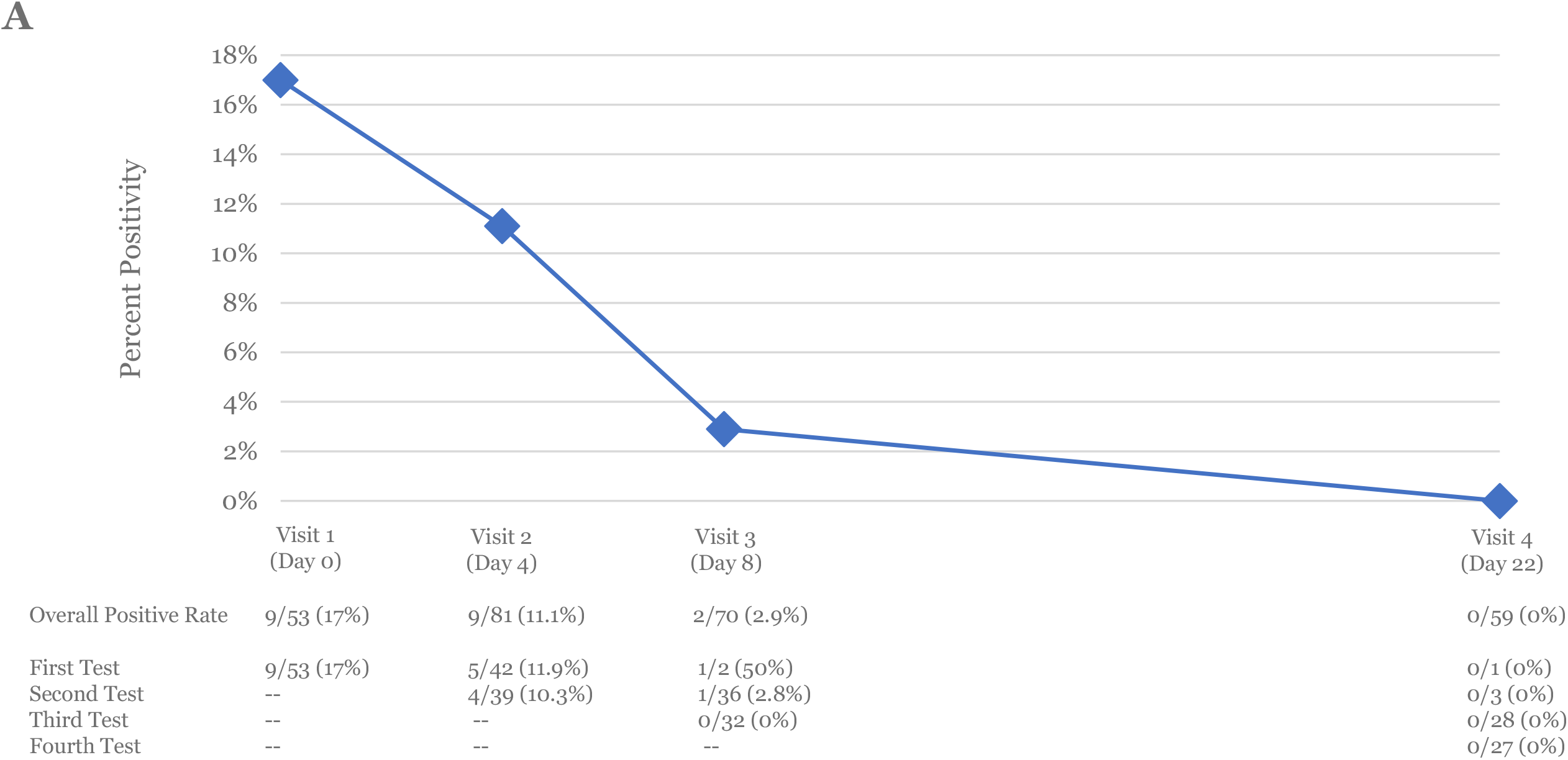
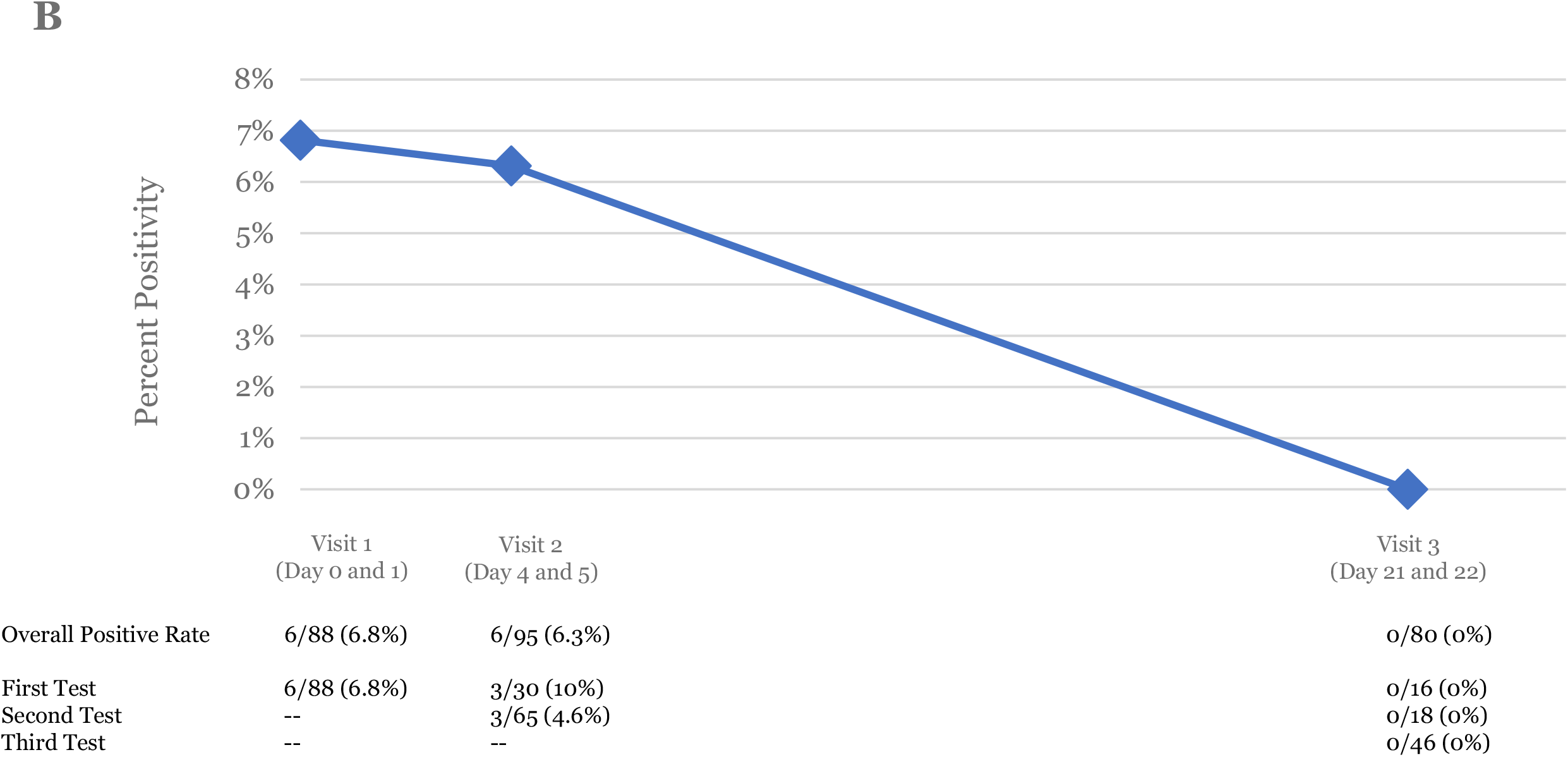
Evolution outbreaks serendipitously found by the study team for (A) Business A and (B) Business B.

Outbreak progression of Business C (manufacturing and supplier sector) is shown in Figure e1. On initial testing, 6/113 (5.3%) were positive, compared to 4/463 (0.9%) on our second visit, and 0/131 on our final visit; retesting acceptance was 100%. Up to March 26, 2021 (2 weeks after our final visit) no new infections were reported. Business D and E were in the meat processing sector and referred by public health due to presumed outbreaks. Among 47 consenting employees at Business D none had SARS-CoV-2 infections, so we did not return for further testing. We tested 136 consenting employees at Business E and 6 (4.4%) were positive. We were unable to schedule a return visit before the end of the study; an additional two persons developed symptoms and tested positive up to March 26, 2021 (three weeks after our visit).

### Interpretation

In this study, we found that systematic, onsite sampling of asymptomatic essential workers with saline gargle was acceptable, safe, and of modest cost. The yield of a first test among asymptomatic employees at businesses with at least two known SARS-CoV-2 infections among workers (an outbreak) was 11-fold higher (4.4% vs. 0.4% positive) compared to businesses not experiencing an outbreak. Among businesses experiencing an outbreak, isolation of positive persons and repeated testing of negative persons over a 2-3 week period detected new infections and appeared to stop transmission; this benefit persisted for at least 2 weeks from our last visit.

Workplaces have been a substantial source of transmission during the COVID-19 pandemic.(14,22) Without systematic testing, infection detection relies on symptomatic workers presenting for testing. Yet, we observed ongoing transmission among asymptomatic workers several weeks after symptomatic infections had been detected. This suggests more proactive testing strategies are required, however with an estimated 2.6 million essential workers in Canada at high risk of SARS-CoV-2 exposure,(23) testing programs will need to prioritize who and where to test.

Benefits of testing to curb transmission appear greatest when used repeatedly in businesses with a few infections. Repeat testing among businesses experiencing outbreaks serves multiple purposes. Most importantly, repeating visits every 4-5 days at the onset of an outbreak identifies people who may have been recently infected but initially tested negative. Repeat visits also permit testing of employees who could not be tested during previous visits. A final test a few weeks later confirms no new infections have arisen. When compared to costs to the businesses of lost production and to the workers of lost wages associated with shuttering businesses, the costs of repeated systematic testing are inexpensive and would seem to be a significantly better approach.

Saline gargle for RT-PCR is an attractive specimen for onsite sampling.(24) Gargle samples can be self-collected without the presence of a healthcare professional, are stable at room temperature, and are highly acceptable, given the high participation rates on first and repeat testing in this study. Sample materials are inexpensive, and all non-laboratory costs involved in a testing program should be <$10 per person, but RT-PCR costs are non-trivial. Results can potentially be available within hours, but in practice, laboratory turnaround times may cause major delays in getting results(25) if laboratory capacity is inadequate. Further developing laboratory infrastructure or expanding the laboratory network to accredited private laboratories may alleviate these limitations.

Alternatively, developing test capacity for workplace testing outside the laboratory through antigen-based rapid tests could be evaluated.(26) These tests are done at the point-of-care and give same-day results. However, rapid tests have lower sensitivity and specificity than RT-PCR.(27) Their lower sensitivity may be overcome by more frequent testing,(28) but their poorer specificity means positive results need RT-PCR confirmation. A major barrier to their uptake has been logistics including that Health Canada requires some currently available rapid tests be administered and all interpreted by a trained professional.(29) Costs of the tests themselves are $15-$20,(30) but costs associated with administration, interpretation, and personal protective equipment are uncertain, and need for RT-PCR confirmation will vary by epidemiologic setting.

### Limitations

Businesses included in this study were those willing to participate. Though we targeted essential businesses of all types, the acceptance rate may limit generalizability and introduce bias. Three businesses included in this study were referred to the study team by public health, though positive rates among tests done at these businesses (16/889; 1.8%) were like other businesses (37/1737; 2.1%). We did not do sequencing to confirm transmission between participants. However, the identification of new conversions among employees at intervals of 4-5 days strongly suggests ongoing transmission. The absence of new infections at our last visit and for two-weeks post study suggests that testing contributed to ending outbreaks. Finally, though our cost estimation was comprehensive, it did not consider inefficiencies in implementation, such as “dead time” associated with waiting between employees.

## Conclusion

Systematic, onsite gargle sampling of asymptomatic essential workers for RT-PCR was a safe and acceptable method to detect workplace infections, which spares healthcare professionals for other tasks. Repeat testing of all employees appears to be effective in breaking transmission chains. Compared to business closures, systematic testing is inexpensive and likely a much better approach. We believe this strategy improves workplace safety and should be a key component of SARS-CoV-2 preventive efforts.

## Data Availability

Deidentified data may be released in certain circumstances following an approved request with a statistical analysis plan and at the sole discretion of the corresponding author.

## Acknowledgments

The authors would like to thank all persons who took part in the study and all businesses who participated. We would like to thank all members of the research team including, Tanya Branco, Jean-Philippe Brassard, Lise Fougere, Sophie Lachapelle-Chisholm, Gabriella Lockwood, Saeedeh Moayedi-Nia, Ludmila Mouelfi, and Sara Perlman-Arrow. We would like to thank Félix-Antoine Veronneau for his help liaising with community stakeholders and study design. We would also like to thank all stakeholders who contributed to the project and its development and all project monitoring and administrative personnel.

## Author Contributions

Drs. Campbell and Menzies had full access to all the data and take responsibility for the integrity of the data and accuracy of data analysis.

*Concept and Design:* Menzies, Campbell, Uppal, Yansouni.

*Acquisition, Analysis, or Interpretation of Data:* All authors.

*Drafting the Manuscript:* Campbell.

*Critical Revision of the Manuscript for Important Intellectual Content:* All authors.

*Statistical Analysis:* Campbell.

## Conflict of Interest Disclosures

Campbell has provided consulting services for SARS-CoV-2 that are unrelated and outside the submitted work to the COVID-19 Immunity Task Force and The World Bank. All other authors have no conflicts of interest to declare.

## Ethics

This study was approved by the Research Institute of the McGill University Health Centre’s Research Ethics Board (2021-7057); all participants gave explicit, informed consent.

## Funding/Support

This project was supported by the McGill Interdisciplinary Initiative in Infection and Immunity (MI4) through funding provided by the Trottier Family Foundation and Molson Foundation (Grant SCRF-04). Campbell (Award #287869) is funded by a postdoctoral fellowship from the Fonds de Recherche du Québec—Santé. Yansouni holds a “Chercheur-boursier clinicien” career award from the Fonds de Recherche du Québec—Santé.

## Role of the Funder/Sponsor

The funders contributed to initial development of the study question and design. The funders had no role in the conduct of the study; collection, management, analysis, and interpretation of the data; preparation, review, or approval of the manuscript; and decision to submit the manuscript for publication.

## References

1. WHO. COVID-19 Strategic Preparedness and Response Plan [Internet]. Geneva: World Health Organization; 2021 [cited 2021 Apr 23]. Available from: https://www.who.int/publications-detail-redirect/WHO-WHE-2021.02

2. Honein MA. Summary of Guidance for Public Health Strategies to Address High Levels of Community Transmission of SARS-CoV-2 and Related Deaths, December 2020. MMWR Morb Mortal Wkly Rep. 2020;69.

3. Sy KTL, Martinez ME, Rader B, White LF. Socioeconomic Disparities in Subway Use and COVID-19 Outcomes in New York City. American Journal of Epidemiology. 2020 Dec 29;(kwaa277).

4. Mutambudzi M, Niedzwiedz C, Macdonald EB, Leyland A, Mair F, Anderson J, et al. Occupation and risk of severe COVID-19: prospective cohort study of 120 075 UK Biobank participants. Occup Environ Med. 2021 May 1;78(5):307–14.

5. Public Health Agency of Canada. Individual and community-based measures to mitigate the spread of COVID-19 in Canada [Internet]. Ottawa (ON): Government of Canada; 2021 [cited 2021 Apr 23]. Available from: https://www.canada.ca/en/public-health/services/diseases/2019-novel-coronavirus-infection/health-professionals/public-health-measures-mitigate-covid-19.html#a3

6. Ontario Ministry of Health. COVID-19 Reference Document for Symptoms [Internet]. Toronto (ON): Government of Ontario; 2020 [cited 2021 Apr 23]. Available from: https://www.health.gov.on.ca/en/pro/programs/publichealth/coronavirus/docs/2019_reference_doc_symptoms.pdf

7. Yanes-Lane M, Winters N, Fregonese F, Bastos M, Perlman-Arrow S, Campbell JR, et al. Proportion of asymptomatic infection among COVID-19 positive persons and their transmission potential: A systematic review and meta-analysis. PLOS ONE. 2020 Nov 3;15(11):e0241536.

8. Li F, Li Y-Y, Liu M-J, Fang L-Q, Dean NE, Wong GWK, et al. Household transmission of SARS-CoV-2 and risk factors for susceptibility and infectivity in Wuhan: a retrospective observational study. The Lancet Infectious Diseases. 2021 May 1;21(5):617–28.

9. McElfish PA, Purvis R, James LP, Willis DE, Andersen JA. Perceived Barriers to COVID-19 Testing. Int J Environ Res Public Health. 2021 Feb 25;18(5).

10. Sethuraman N, Jeremiah SS, Ryo A. Interpreting Diagnostic Tests for SARS-CoV-2. JAMA. 2020 May 6;323(2):2249–51.

11. Alberta Health Services Infection Prevention & Control. IPC Recommendations PPE Table for Assessment Centres during COVID-19. Alberta (Canada): Alberta Health Services; 2021.

12. Frazee BW, Rodríguez-Hoces de la Guardia A, Alter H, Chen CG, Fuentes EL, Holzer AK, et al. Accuracy and Discomfort of Different Types of Intranasal Specimen Collection Methods for Molecular Influenza Testing in Emergency Department Patients. Ann Emerg Med. 2018;71(4):509–17.

13. Bastos ML, Perlman-Arrow S, Menzies D, Campbell JR. The Sensitivity and Costs of Testing for SARS-CoV-2 Infection With Saliva Versus Nasopharyngeal Swabs. Ann Intern Med. 2021 Jan 12;174(4):501–10.

14. Sante Montreal. Current COVID-19 Situation in Montreal [Internet]. 2021 [cited 2021 Apr 20]. Available from: https://santemontreal.qc.ca/en/public/coronavirus-covid-19/situation-of-the-coronavirus-covid-19-in-montreal/

15. INSPQ. COVID-19 Data in Quebec [Internet]. INSPQ. 2021 [cited 2021 May 5]. Available from: https://www.inspq.qc.ca/covid-19/donnees

16. Tourisme Montreal. Current Situation in Montreal [Internet]. 2021 [cited 2021 Apr 20]. Available from: https://www.mtl.org/en/covid-19

17. Rowe DJ. What is considered an essential service in Quebec? CTV [Internet]. 2020 Dec 26 [cited 2021 Apr 30]; Available from: https://montreal.ctvnews.ca/what-is-considered-an-essential-service-in-quebec-here-s-a-list-1.5245449

18. MSSS. Répertoire québécois et système de mesure des procédures de biologie médicale. Quebec: Ministère de la Santé et des Services Sociaux; 2021.

19. MSSS. Répertoire québécois et système de mesure des procédures de biologie médicale (Annex). Quebec: Ministère de la Santé et des Services Sociaux; 2021.

20. Gilmour AR, Anderson RD, Rae AL. The analysis of binomial data by a generalized linear mixed model. Biometrika. 1985 Dec 1;72(3):593–9.

21. Abrahantes JC, Aerts M. A solution to separation for clustered binary data. Statistical Modelling. 2012 Feb 1;12(1):3–27.

22. Toronto Public Health. COVID-19:Status of Cases in Toronto [Internet]. City of Toronto. City of Toronto; 2020 [cited 2021 Apr 26]. Available from: https://www.toronto.ca/home/covid-19/covid-19-latest-city-of-toronto-news/covid-19-status-of-cases-in-toronto/

23. Campbell JR, Uppal A, Oxlade O, Fregonese F, Bastos ML, Lan Z, et al. Active testing of groups at increased risk of acquiring SARS-CoV-2 in Canada: costs and human resource needs. CMAJ. 2020 Oct 5;192(40):E1146–55.

24. Goldfarb DM, Tilley P, Al-Rawahi GN, Srigley JA, Ford G, Pedersen H, et al. Self-Collected Saline Gargle Samples as an Alternative to Health Care Worker-Collected Nasopharyngeal Swabs for COVID-19 Diagnosis in Outpatients. J Clin Microbiol. 2021 Mar 19;59(4).

25. Warnica R. Inside Ontario’s overwhelmed labs: How lingering issues and mistakes caused massive COVID-19 testing backlog. National Post [Internet]. 2020 [cited 2021 Apr 26]; Available from: https://nationalpost.com/news/canada/inside-ontarios-overwhelmed-labs-how-lingering-issues-and-mistakes-caused-massive-covid-19-testing-backlog

26. Government of Ontario. COVID-19 Rapid Screening Pilot Program Launching in Waterloo Region [Internet]. news.ontario.ca. 2021 [cited 2021 May 5]. Available from: https://news.ontario.ca/en/release/61100/covid-19-rapid-screening-pilot-program-launching-in-waterloo-region

27. Peeling RW, Olliaro PL, Boeras DI, Fongwen N. Scaling up COVID-19 rapid antigen tests: promises and challenges. The Lancet Infectious Diseases. 2021 Feb 23;epub ahead of print.

28. Larremore DB, Wilder B, Lester E, Shehata S, Burke JM, Hay JA, et al. Test sensitivity is secondary to frequency and turnaround time for COVID-19 screening. Sci Adv. 2021 Jan;7(1).

29. Health Canada. COVID-19 testing devices: Authorized medical devices [Internet]. aem. 2021 [cited 2021 Apr 30]. Available from: https://www.canada.ca/en/health-canada/services/drugs-health-products/covid19-industry/medical-devices/authorized/list.html

30. Rapid Test & Trace Canada [Internet]. 2021 [cited 2021 May 5]. Available from: https://rapidtestandtrace.ca/

