## Appendix for "Systematic Testing for SARS-CoV-2 Infection Among Essential Workers in Montréal, Canada: A Prospective Observational and Cost Assessment Study"

**Appendix 1 (as supplied by the authors): Systematic Testing for SARS-CoV-2 Infection Among Essential Workers in Montréal, Canada: A Prospective Observational and Cost Assessment Study**

### Additional Information

#### Sampling Protocol

**Materials**

*Base Materials:*

- Sanitizing materials (wipes, alcohol spray, towels, rags, etc.)
- Barrier between participant and person collecting sample
- 1 tube rack
- 1 bottle of hand sanitizer with pump
- 1 trash can
- Table and chairs as necessary

*Recurrent Materials (per person):*

- 1 pair of gloves
- 2 masks (if necessary – one for participant, one for person collecting sample [this can be used across multiple participants])
- 2 biohazard specimen bags with an outside pocket
- 5ml ampoule of saline solution
- 10ml sterile tube
- 30ml collection cup
- Participant label for the sterile tube

**Procedures**

*Preparation*

1. Set up the barrier on a recently sanitized table with the tube rack on the participant side of the barrier.
2. Prior to each participant arriving, in the tube rack place the 5ml ampoule of saline solution and 10ml sterile tube. Next to the tube rack place the 30ml collection cup.
3. Place the hand sanitizer on the participant's side of the barrier and place the garbage can next to the participant's seat.
4. On the side of the barrier for the person collecting the sample, place 2 biohazard specimen bags, a participant label for the sterile tube, and any required paperwork that will need to be completed for the laboratory.

*Sample Collection*

1. The person collecting the sample should put on a fresh pair of gloves and be wearing a surgical mask meeting minimum Health Canada requirements.
2. When the participant arrives, instruct them to have a seat and use the hand sanitizer. Offer them a mask should they not have one.
3. Collect the required laboratory information from the participant and any information required for the participant label (e.g., name and date of birth). Confirm the information with the participant.
4. Introduce the sample collection procedure to the participant (using steps 5 through 11 below as a guide).
5. Instruct the participant to open the saline ampoule and empty the contents into the collection cup. Discard the ampoule in the garbage can.
6. Instruct the participant to temporarily remove their mask to place the saline solution in their mouth. Remind the participant to put their mask on after.
7. Instruct the participant to follow these steps:
    a. Swish the saline solution in their mouth for 5 seconds.
    b. Gargle the saline solution in their throat for 5 seconds.
    c. Repeat (a) and (b) one more time.
8. When the steps are completed, instruct the participant to temporarily remove their mask and spit the contents of their mouth back into the collection cup. Remind the participant to put their mask on after.
9. Instruct the participant to open the sterile tube.

10. Instruct the participant to pinch the collection cup to create a pseudo-funnel to empty the contents of the collection cup into the sterile tube.
11. Instruct the participant to discard the collection cup, close the sterile tube, and place the sterile tube back in the rack.
12. Instruct the participant to sanitize their hands. They are now free to leave.

*Sample Storage*

1. When the participant has left, sanitize their side of the table.
2. Sanitize the outside of the sterile tube and ensure the tube cap has been placed on securely.
3. Attach the participant label to the sanitized tube.
4. Place the tube inside one biohazard bag; remove the air and seal securely.
5. Place the biohazard bag with the tube into the other biohazard bag; remove the air and seal securely.
6. To the outside pouch of the biohazard bag, include any documentation (e.g., requisition form) required by the laboratory.
7. Store the sample in a secure place; the sample is stable at room temperature for 7 days.

#### Cost Assessment Details

We conceptualized cost considerations across six distinct steps:

1. Training of staff
2. Scheduling and coordination of visits
3. Sample collection
4. Sample transport
5. Laboratory reverse-transcription polymerase chain reaction (RT-PCR)
6. Communication of results

We considered three different staff roles:

- Coordinator: $30 per hour (inclusive of benefits)
- Mobile Team Member: $23 per hour (inclusive of benefits)
- Administrative: $20 per hour (inclusive of benefits)

We assumed two mobile team members could sample 100 persons per day or 26,000 persons per year.

<u>Training of Staff</u>

Based on our experience with the study, we estimated that training outside of a research context could be done over 2 days (7h/day) by a coordinator. We assumed this training would need to be done annually. This training would make use of sample collection materials and personal protective equipment. Each day, we assumed each mobile team member would use the equivalent of materials required to sample 10 persons (e.g., each training day, material costs equivalent to sampling 20 persons). We included a cost of $15 per person, per day for food and refreshments.

<u>Scheduling and Coordination of Visits</u>

Based on our experience in the study, we estimated that scheduling and coordination of visits would require 2h/business by administrative staff. In practice, coordination would include discussion with the business regarding accessibility, location for sampling stations, and other business-specific information. Scheduling would be done in conjunction with the business to ensure processes run smoothly (e.g., ensure employees come every 6 minutes to be sampled to reduce crowding).

<u>Sample Collection</u>

Based on our experience in the study, we estimated that it would take 25 minutes to set up sample stations in the business—this includes unloading the materials, bringing them to the sampling location, and preparing the station for the first participant.

We considered capital purchases required for each station (e.g., chairs, tables, plexiglass, tube racks, sample coolers, bins for materials, trash bins, stationery, dollies to move materials, computer/tablet required for data collection for lab). We assumed these would all need to be replaced annually.

To estimate time associated with sampling participants, measuring time on the initial test was not considered to be representative of how long it would take in practice. This was because data collection for the research study was done throughout the process with the participant and not clearly separated from sample collection. Therefore, we estimated time during re-testing of participants where re-consent was not necessary nor was questionnaire completion. We measured how many participants were sampled by the mobile team over a day in a well coordinated business. One mobile team member re-tested 100 participants over 7h of working time (average time of 4.2 minutes per participant). This was communicated as “not sustainable” by the mobile team member day-in-and-day-out and thus in practice we estimated a more realistic time would be 6 minutes per participant.

For costs associated with sampling, we assumed that each mobile team member would require 4 surgical masks per day and 2 gloves per participant sampled (eye protection and gowns were not required given samples were self-collected behind a physical barrier). To estimate cost associated with sanitizing materials we used the actual costs incurred over the study. Taking into consideration the remaining sanitizing materials at the end of the study, total costs were ~$800. We assumed one trash bag would be required per business. Sample materials included 2 biohazard bags, a tube label, a 5ml saline ampoule, a 10ml sample tube, and a 30ml collection cup.

Based on our experience in the study, we estimated that it would take 20 minutes to tear down sample stations in the business, sanitize surfaces, and pack materials into the vehicle.

Sample Transport

We used data from the study and estimated the transport time between businesses was 10 minutes and end of day transport to the laboratory was 30 minutes. We used mileage from vehicles during the study and estimated each vehicle was driven ~60km each day. We used Government of Canada[1] mileage reimbursement to estimate costs associated with each travel.

Laboratory RT-PCR

We used the Québec government reimbursement price for RT-PCR analyses related to SARS-CoV-2. This price includes cost of sample accessioning, analysis, and sending of all results to the requesting provider and positive results to public health.

Communication of Results

During our study, we estimated the time to receive results, organize them, and send a text/email to negative participants was just under 5 minutes per person by administrative staff. For persons testing positive, phone calls took approximately 10 minutes and done by a nurse. Only 1/50 tests were positive in our study. If a nurse was paid $30 per hour, each call to a positive participant would be $5. The cost for administrative personnel for negative participants would be $1.58. When combined, *on balance* time associated with communication of results is equivalent to the cost of administrative personnel taking 5 minutes per person. If positive rates doubled (1/25) or quintupled (1/10) then the cost would be equivalent to 5.2 minutes and 5.8 minutes, respectively, of administrative staff time per person. We felt this was negligible and did not consider varying costs.

---

[1] https://www.canada.ca/en/revenue-agency/services/tax/businesses/topics/payroll/benefits-allowances/automobile/automobile-motor-vehicle-allowances/automobile-allowance-rates.html

#### Sample Size

Our initial sample size calculation was based on SARS-CoV-2 rates seen in Montréal during the late fall 2020 and previous reported experience of SARS-CoV-2 positivity among asymptomatic essential workers in Calgary (1.6%).[2] We determined a target sample size of 5% among essential workers in Montréal-North could be realistic given the state of the pandemic and this previous experience. We used our previous work estimating the number of essential workers across Canada to estimate that there was likely around 5000 essential workers in Montréal-North working in 30 essential businesses with >20 employees and 200 essential businesses with ≤20 employees. Considering this finite sample size, using a binomial estimator, an intraclass correlation coefficient of 0.3 between businesses, and an absolute precision of ±2% around the 5% target, we estimated 2,589 participants would be required to estimate a 5% prevalence with a 95% confidence interval of 3% to 7%. When our study began, the situation in Montréal was substantially different, with several lockdown restrictions in place and a province-wide curfew, which likely impacted community prevalence and our initial estimates of prevalence from late fall 2020.

[2] https://globalnews.ca/news/6961600/coronavirus-calgary-asymptomatic-testing-results/

### Tables

#### Table e1. Questionnaire Elements

| Variable / Question | Possible Responses (If Applicable) |
|---|---|
| Personal Information (for Lab/Public Health) | |
| Today's Date | |
| Business Name | |
| Participant ID | |
| Last and First Name | |
| Date of Birth | |
| Address | |
| Telephone Number | |
| Email | |
| Health Insurance Number | |
| Demographic Information | |
| Age | |
| Sex | Male, Female |
| Ethnicity | Asian, Black, Caucasian, Hispanic, Indigenous, Mixed, Other |
| Employment Status | Full-Time, Part-Time, Occasional |
| Clinical Information | |
| Health Conditions and Habits | Hypertension, Diabetes, Chronic Respiratory Conditions, Heart Disease, Current or Past Cigarette Smoking, Other |
| Travel Outside of Greater Montréal | Yes (Where), No |
| Travel Outside of Québec | Yes (Where), No |
| History of COVID-19 Contact | Yes, No |
| Feeling Today | Fine, Not My Best, Quite Ill |
| Symptoms Today | Fever, Dry Cough, Fatigue, Aches and Pains, Sore Throat, Diarrhea, Conjunctivitis, Headache, Anosmia or Loss of Taste, Rash, Difficulty Breathing or Shortness of Breath, Chest Pain or Pressure, Loss of Speech or Movement, Other |
| Symptoms in Previous Two Weeks | Fever, Dry Cough, Fatigue, Aches and Pains, Sore Throat, Diarrhea, Conjunctivitis, Headache, Anosmia or Loss of Taste, Rash, Difficulty Breathing or Shortness of Breath, Chest Pain or Pressure, Loss of Speech or Movement, Other |
| Previous SARS-CoV-2 Test | Yes, No |
| Date of Previous SARS-CoV-2 Test | |
| Result of Previous SARS-CoV-2 Test | |

#### Table e2. Detailed Cost Components

| Cost Estimate | Additional Details | Cost per Unit* | Cost per Person Sampled† |
|---|---|---|---|
| **Training** | | $2,058.00 | $0.04 |
| Mobile Team and Coordinator Time | Four mobile team members and one coordinator for 2 days | $1,708.00 | -- |
| Food | $15 per person per day | $150.00 | -- |
| Sample Materials | For 10 persons per mobile team member per day | $139.20 | -- |
| PPE | For all attendees, 10 sets | $60.80 | -- |
| **Scheduling** | | $40.00 | $0.80 |
| Administrative Personnel Time | 2 hours per business | $40.00 | -- |
| **Sample Collection** | | $7,631.92 | $5.62 |
| *Capital Purchases* | | $3,376.59 | $0.18 |
| Chairs | Four chairs per team | $165.28 | $0.01 |
| Tables | Two tables per team | $137.90 | $0.01 |
| Plexiglass | Plexiglass barriers per team (2 small, 1 large) | $467.11 | $0.02 |
| Cooler | One sample cooler for transport | $34.00 | <$0.01 |
| Tube Racks | Two tube racks per team | $40.00 | <$0.01 |
| Bins for Materials | Eight bins per team | $90.40 | <$0.01 |
| Trash Bins | Two per team | $94.90 | <$0.01 |
| Stationery | Pens, paper, printing lab forms, etc. | $1,000.000 | $0.04 |
| Dollies | One per team | $147.00 | $0.01 |
| Computer/Tablets | Two per team | $1,200 | $0.09 |
| *Recurrent Purchases* | | | $2.45 |
| Mask | Four per day per team member | $1.60 | $0.02 |
| Gloves | Two gloves per person | $0.36 | $0.36 |
| Biohazard Bags | Two per person | $0.24 | $0.24 |
| Sample Tube | One per person | $0.51 | $0.51 |
| Saline Ampoule | One per person | $0.83 | $0.83 |
| Collection Cup | One per person | $0.06 | $0.06 |
| Sample Label | One per person | $0.10 | $0.10 |
| Sanitizing Materials | Sanitizer, alcohol sprays, disinfectant wipes, paper towels (realized cost during study) | $800.00 | $0.30 |
| Trash Bag | Two per business | $1.44 | $0.03 |
| *Mobile Team Personnel Time* | | $36.80 | $2.99 |
| Setup | 25 minutes per member, per business | $19.17 | $0.38 |
| Tear Down | 20 minutes per member, per business | $15.33 | $0.31 |
| Sample Collection | 6 minutes per member, per person | $2.30 | $2.30 |
| **Transport** | | $54.57 | $0.54 |
| Mobile Team Personnel Time | 10 minutes of driving between businesses and 30 minutes for one member to drop off samples | $19.17 | $0.19 |
| Fuel & Mileage | 60km per day at $0.59 per km | $35.40 | $0.35 |
| **Laboratory RT-PCR** | | $34.00 | $34.00 |
| Reimbursement cost in Québec | Inclusive of accessioning and sending results back to person/facility ordering the test | $34.00 | -- |
| **Communication of Results** | | $1.67 | $1.67 |
| Administrative Personnel Time | 5 minutes—on balance—per person | $1.67 | -- |
| **OVERALL COST PER PERSON** | -- | -- | **$42.67** |

*See earlier in appendix for additional details, salaries, and assumptions
†Assuming two 50-person businesses visited per day by two mobile team members and 26,000 persons sampled per year

#### Table e3. Positive Results Among Participants by Test Number

| Parameter | Non-Outbreak Settings | Outbreak Settings | | | | |
|---|---|---|---|---|---|---|
| **Businesses** | 64* | Business A† | Business B† | Business C†† | Business D†† | Business E†† |
| **1st Test** | 5/1211 (0.4%) | 15/98 (15.3%) | 9/134 (6.7%) | 0/47 (0%) | 6/136 (4.4%) | 10/502 (2.0%) |
| **2nd Test** | -- | 5/78 (6.4%) | 3/83 (3.6%) | --- | --- | 0/189 (0%) |
| **3rd Test** | -- | 0/60 (0%) | 0/46 (0%) | --- | --- | 0/15 (0%) |
| **4th Test** | -- | 0/27 (0%) | --- | --- | --- | --- |
| **Overall** | **5/1211 (0.4%)** | **20/263 (7.6%)** | **12/232 (5.2%)** | **0/47 (0%)** | **6/136 (4.4%)** | **10/706 (1.4%)** |

*Includes a separate building not experiencing an outbreak in a business with an outbreak.
†Serendipitously found by the study team
††Referred to the study team by public health.

#### Table e4. Crude Odds Ratios for Factors Potentially Associated with a Positive Test

| Factor | Number of Participants (N=2128) | Tests Positive (%) | Crude Odds Ratio† (95% CI) |
|---|---|---|---|
| **Age (per 1-year increase)** | -- | -- | 1.0 (0.98 to 1.03) |
| **Sex** | | | |
| Female | 808 | 9 (1.1%) | 1.0 (reference) |
| Male | 1320 | 44 (3.3%) | 2.4 (1.1 to 5.5) |
| **Self-Reported Ethnicity** | | | |
| Caucasian | 926 | 4 (0.4%) | 1.0 (reference) |
| Non-Caucasian | 1202 | 49 (4.1%) | 4.0 (1.5 to 10.6) |
| **Health Factor** | | | |
| None Reported | 1681 | 45 (2.7%) | 1.0 (reference) |
| Any Reported* | 447 | 8 (1.8%) | 0.7 (0.3 to 1.6) |
| **Smoking History** | | | |
| Never Smoker | 1668 | 46 (2.8%) | 1.0 (reference) |
| Current or Previous Smoker | 460 | 7 (1.5%) | 0.6 (0.3 to 1.4) |
| **Feeling on Day of Testing**** | | | |
| "Fine, Same as Any Other Day" | 2098 | 51 (2.4%) | 1.0 (reference) |
| "Not My Best Today" | 30 | 2 (6.7%) | 2.1 (0.4 to 10.0) |
| **Previous SARS-CoV-2 Test History***** | | | |
| Never Previously Tested for SARS-CoV-2 | 1155 | 42 (3.6%) | 1.0 (reference) |
| Previously Tested for SARS-CoV-2 and Always Negative | 882 | 9 (1.0%) | 0.3 (0.1 to 0.7) |
| Previously Tested for SARS-CoV-2 and Positive | 91 | 2 (2.2%) | 0.7 (0.2 to 3.0) |
| **Business Size** | | | |
| ≤50 Participants | 895 | 4 (0.4%) | 1.0 (reference) |
| >50 Participants | 1233 | 49 (4.0%) | 10.4 (0.7 to 146.5) |

Abbreviations: SARS-CoV-2, severe acute respiratory syndrome coronavirus2
†The model accounts for clustering by business and sector
*includes hypertension, diabetes, chronic respiratory conditions (e.g., asthma), heart disease, or participant reported conditions
**refers to reporting on electronic questionnaire (not to study team), which was reviewed post-visit
***considers only tests done prior to study enrolment

#### Table e5. Sensitivity Analysis of Costs Per Person Sampled

| Cost Estimates | Cost Per Person Sampled ($ CAD) | | | |
|---|---|---|---|---|
| | Two 50-person businesses daily | Eight 5-person businesses daily | Four 20-person businesses daily | One 120-person business daily |
| **TOTAL COST** | **$42.67** | **$58.33** | **$45.28** | **$41.62** |
| **TOTAL LABORATORY COST** | **$34.00** | **$34.00** | **$34.00** | **$34.00** |
| **TOTAL NON-LABORATORY COST** | **$8.67** | **$24.33** | **$11.28** | **$7.62** |
| **Training of Mobile Team** | **$0.04** | **$0.04** | **$0.04** | **$0.04** |
| Coordinator & Mobile Team Personnel | $0.03 | $0.03 | $0.03 | $0.03 |
| Materials & Refreshments | $0.01 | $0.01 | $0.01 | $0.01 |
| **Scheduling Businesses** | **$0.80** | **$8.00** | **$2.00** | **$0.33** |
| Administrative Personnel | $0.80 | $8.00 | $2.00 | $0.33 |
| **Sample Collection** | **$5.62** | **$12.11** | **$6.70** | **$5.19** |
| Mobile Team Personnel | $2.99 | $9.20 | $4.03 | $2.59 |
| Sanitizing Materials | $0.33 | $0.59 | $0.38 | $0.32 |
| Personal Protective Equipment | $0.38 | $0.40 | $0.38 | $0.37 |
| Sample Collection Materials | $1.74 | $1.74 | $1.74 | $1.74 |
| Capital Purchases | $0.18 | $0.18 | $0.18 | $0.18 |
| **Sample Transport** | **$0.54** | **$2.51** | **$0.87** | **$0.39** |
| Mobile Team Personnel | $0.19 | $1.62 | $0.43 | $0.09 |
| Vehicle & Fuel | $0.35 | $0.89 | $0.44 | $0.30 |
| **Communicating Results** | **$1.67** | **$1.67** | **$1.67** | **$1.67** |
| Administrative Personnel | $1.67 | $1.67 | $1.67 | $1.67 |

### Figures

#### Figure e1. Outbreak Progression at Business C

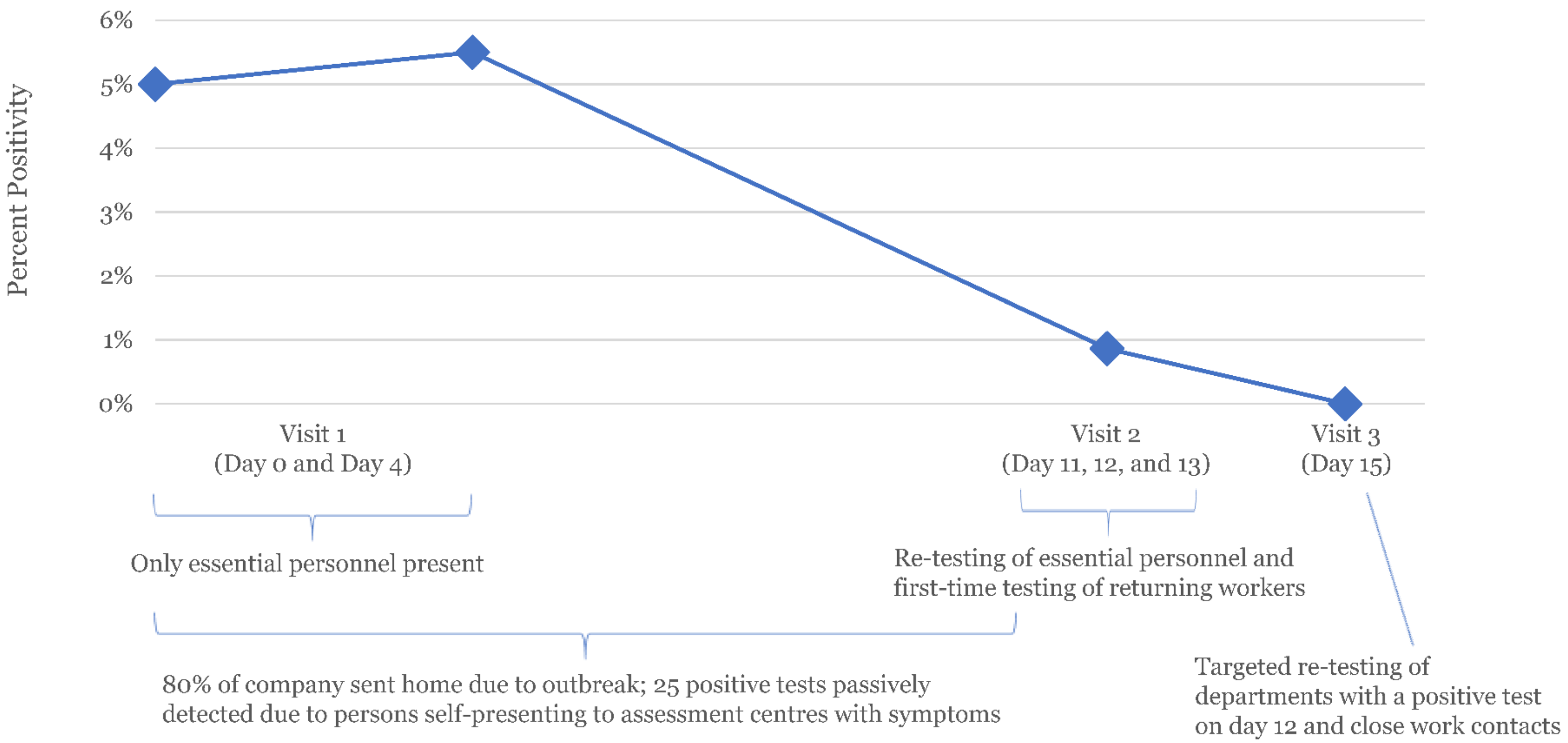

| | | | | |
|---|---|---|---|---|
| Overall Positive Rate | 3/58 (5.2%) | 3/55 (5.5%) | 4/463 (0.9%) | 0/130 (0%) |
| First Test | 3/58 (5.2%) | 3/55 (5.5%) | 4/357 (1.1%) | 0/32 (0%) |
| Second Test | -- | -- | 0/106 (0%) | 0/83 (0%) |
| Third Test | -- | -- | -- | 0/15 (0%) |

*Caption:* Business C was referred by public health and was in the manufacturing and supplier sector. It had temporarily reduced staffing by 80% to combat an ongoing outbreak. Due to staggered schedules, our initial visit was done over two visits, 4 days apart—we found 6/113 (5.3%) participants tested positive. All employees returned to work on day 11. Over three consecutive days we offered testing to all returning employees and all persons initially negative. On the first day, none of the 106 participants being re-tested were positive (100% testing uptake). None of the returning employees consenting to testing on day one (105 participants) and day three (121 participants) were positive. However, on the second day, 4/131 (3.1%) of participants receiving their first test were positive. We returned to the business three days later to conduct targeted re-testing among departmental and other close contacts who were previously negative—none of the 130 tests were positive (100% testing uptake). Up to March 26, 2021 (two weeks after study end) no new infections have been reported.
